# Ultrasound applications in the treatment of major depressive disorder (MDD): A systematic review of techniques and therapeutic potentials in clinical trials and animal model studies

**DOI:** 10.1101/2025.01.23.25320960

**Authors:** Gansheng Tan, Hong Chen, Eric C. Leuthardt

## Abstract

**Objective:** Major depressive disorder (MDD) is a debilitating condition that inflicts significant personal and economic burdens and affects around 8% of the US population. Approximately 30% of patients with MDD do not respond to conventional antidepressant and psychotherapeutic treatments. Current treatment options for refractory MDD include transcranial magnetic stimulation (TMS) and invasive surgical procedures such as surgical ablation, vagus nerve stimulation, and deep brain stimulation. In this context, therapeutic ultrasound emerges as a promising alternative for treating refractory MDD, which has the unique advantage of combining non-invasiveness with selective targeting. Over the past 10 years, there has been a growth in focused ultrasound research, leading to an exponential increase in interest in the technology. To support the future development of ultrasound for treating MDD, we conducted a systematic review following Preferred Reporting Items for Systematic Review and Meta-Analyses (PRISMA) guidelines.

**Materials and Methods:** We identified 86 relevant studies from 1975 through June 2025. Our inclusion criteria were peer-reviewed prospective cohort studies, case-control studies, and randomized controlled trials that report ultrasound efficacy for treating depression in humans or depressive-like behaviors in animal models (PROSPERO registration number: CRD42024626093). 23 studies met all inclusion criteria. We summarized ultrasonic techniques for treating depression and their efficacy.

**Results:** Two focused ultrasound (FUS) techniques used to treat depression include magnetic resonance-guided focused ultrasound (MRgFUS) for capsulotomy and low-intensity focused ultrasound (LIFUS) neuromodulation. MRgFUS capsulotomy results in permanent lesioning, whereas LIFUS is non-lesional and thought to have temporary effects. In human trials, the response rate (≥50% improvement in depression score from baseline) for MRgFUS capsulotomy and LIFUS neuromodulation were 53.85% and 69.2%, respectively. The odds ratio for LIFUS was 2.8. In addition, LIFUS neuromodulation had a large effect (|Cohen’s d| > 0.8) on reducing standard depression scale scores in humans or resolving depressive-like behaviors in rodents. The certainty of evidence is moderate for human trials and low for rodent models. MRgFUS capsulotomy had inconsistent lesioning success and a limited response rate, while LIFUS neuromodulation lacked systematic exploration of the parameter space and clear delineation of the underlying mechanisms. Future work should refine patient selection for MRgFUS capsulotomy and optimize the parameters for individualized functional targeting.

**Conclusions:** LIFUS neuromodulation achieved a large reduction in depressive behaviors in both rodent models and human trials. We conclude that LIFUS neuromodulation is a promising, noninvasive option for treating refractory MDD.

## Introduction

Major depressive disorder (MDD) is a severe form of mood disorder characterized by persistent feelings of sadness or anhedonia (inability to experience pleasure). It is often accompanied by feelings of guilt or worthlessness, changes in appetite, sleep disturbances, fatigue, and cognitive impairments. Approximately 21 million adults in the United States (8.3% of all adults) had at least one major depressive episode in 2021^1^. The etiology of MDD is believed to be multifactorial, including genetic, environmental, and psychosocial factors. Widely accepted theories of MDD pathogenesis include deficiencies in monoamine neurotransmitters, the hypothalamic–pituitary–adrenal (HPA) axis dysfunction, and increased inflammatory signaling^2–4^. Recent neuroimaging studies suggest that network dysfunction plays a role in MDD pathogenesis, providing a theoretical basis for neuromodulation therapy^5,6^. The dorsolateral prefrontal cortex (PFC) and anterior cingulate are key components of mood-regulating circuits. Altered connectivity between these two regions may contribute to depressive pathophysiology^6,7^. Notably, functional connectivity between the subcallosal cingulate, a subregion of the anterior cingulate, and the dorsolateral PFC has been shown to predict treatment response to PFC stimulation^7,8^. The serotonin theory of depression, a part of the monoaminergic theory of depression, posits that reduced serotonin activity contributes to depressive symptoms^9^. This theory was first suggested in the 1960s, which has promoted the use of antidepressants, particularly selective serotonin reuptake inhibitors, as standard treatments for MDD^10^. The response rate to a first antidepressant medication typically ranges from 40% to 60%, compared to 20%–40% for a placebo^11^. A large-scale trial showed that nearly 80% of the outpatients treated with antidepressants had chronic or recurrent major depression^12^, and approximately 30% of patients with refractory MDD resisted further treatments after failed treatment attempts^13^. For these patients, invasive neurosurgical procedures such as bilateral anterior capsulotomy, bilateral anterior cingulotomy, subcaudate tractotomy, vagus nerve stimulation, and deep brain stimulation are sometimes pursued. These surgical approaches aim to disrupt the aberrant brain networks implicated in MDD and have shown therapeutic potential, although they yield variable clinical outcomes and carry surgical risks such as infection and hemorrhage^14,15^. In 2008, the US Food and Drug Administration (FDA) approved the use of transcranial magnetic stimulation (TMS) for MDD, which works by non-invasively delivering magnetic pulses to the brain. The standard TMS treatment protocol typically requires daily sessions over several weeks^16^. Existing systematic reviews of TMS for depression that included studies conducted before 2000 reported response rates around 29%^17–19^. A more recent systematic review, restricted to studies published after 2000, reported a response rate closer to 40%, compared to 14% in the sham control^16^. Other neuromodulation techniques for treating depression include electroconvulsive therapy, transcutaneous vagus nerve stimulation, and transcranial direct current stimulation^20–23^.

In recent years, there has been increasing interest in using ultrasound for treating depression, largely because it can reach deeper brain targets implicated in MDD and can be precisely focused to target specific brain regions. Ultrasound technology can be categorized into diagnostic ultrasound and therapeutic ultrasound. This review focuses specifically on therapeutic ultrasound. Therapeutic ultrasound directs the ultrasound (i.e., mechanical waves with frequencies greater than 20 kHz) to the target through focused ultrasound transducers. Effective ultrasound transmission requires a coupling medium (e.g., ultrasound gel) to be applied between the transducer and tissue because absorption and reflection occur at interfaces between materials of different acoustic impedance. Technological advances in intraoperative magnetic resonance imaging (MRI), beamforming algorithms for targeting and correcting aberrations caused by the skull, and neuronavigation have facilitated the clinical translation of therapeutic ultrasound^24^. Both high-intensity focused ultrasound (HIFUS) and low-intensity focused ultrasound (LIFUS) are now being explored as therapeutic tools for depression. HIFUS is mostly used for non-invasive tissue ablation, either by inducing coagulative thermal necrosis due to the absorption of ultrasound energy or by causing mechanical ablation through high-tensile pressure-induced cavitation^25^. In contrast, LIFUS is designed to modulate neural activity without irreversible lesions. Neuroimaging studies have demonstrated its ability to modulate brain activity^26–28^. Both animal and human studies evaluating the safety profile of LIFUS found no evidence of tissue damage^26,29–31^. While the exact mechanism mediating LIFUS neuromodulation remains unclear, it is thought to involve acoustic radiation force, cavitation effects (oscillation of gas bubbles within tissues), and thermal effects (for a review of the mechanism, see^32^).

Ultrasound technologies have only recently been explored for the treatment of neuropsychiatric diseases, and earlier reviews mostly provided narrative summaries^33^. Since 2018, a number of clinical trials have begun to investigate these ultrasound technologies in humans^14,34^, and a growing body of studies has generated valuable data on the efficacy of HIFUS and LIFUS for depression^28,35–40^. This review has two main goals. First, we aim to provide an up-to-date summary of techniques and protocols for administering ultrasound in the treatment of depression. This is informative at the current developmental stage, where no standardized intervention protocol exists. Second, we aim to provide preliminary, separate estimates of the efficacy of LIFUS and HIFUS for depression. While the available clinical data are still limited, given the early investigational stage, synthesizing results across small and single-center trials is critical for informing the design of future multi-center, randomized clinical trials.

As with other neuromodulation techniques, animal models, especially rodent models, play a crucial role in the early-stage development of ultrasound application for treating depression. Preclinical studies showed that LIFUS neuromodulation in animal models improved depressive-like behaviors^27,29,30,41–44^. These studies are useful for evaluating safety and efficacy, identifying therapeutic targets, investigating underlying mechanisms, and exploring potential biomarkers, thereby informing subsequent clinical trials. Commonly used depression models include environmentally induced, pharmacological, genetic, and lesion-based paradigms^45–47^. Among these, chronic stress models, such as chronic unpredictable stress and chronic restraint stress, are the most widely used and considered to have good predictive, face, and construct validity^46^. To quantitatively assess the efficacy of antidepressant interventions in rodent models, researchers used standardized behavioral tests^48^. These include the forced swim test and the tail suspension test to measure behavioral despair, the sucrose preference test to assess anhedonia, reduced ability to experience pleasure, and the elevated plus maze to evaluate anxiety-like behavior. While preclinical studies serve as a first step in evaluating novel interventions, caution is warranted when extrapolating these findings to human clinical outcomes because of the unique and complex features of human depression^48^. In this review, we summarize recent advances in ultrasound techniques for depression, including LIFUS neuromodulation and HIFUS thermal ablation. We assess their therapeutic potential in the early translational phase by synthesizing efficacy data separately for clinical and preclinical studies. Additionally, we present the current status of therapeutic ultrasound for depression, identify key challenges, and discuss opportunities for future research and clinical translation.

## Materials and Methods

### Systematic review design

This systematic review was preregistered in PROSPERO (CRD42024626093) and follows the Preferred Reporting Items for Systematic Reviews and Meta-Analyses (PRISMA) reporting guidelines^49^ (**Figure 1**). We included both clinical and preclinical studies, recognizing that each provides complementary insights to the early-stage development of therapeutic ultrasound for depression. Preclinical studies inform safety and target selection, and clinical trials offer initial evidence of feasibility and efficacy in humans.

**Figure 1.**
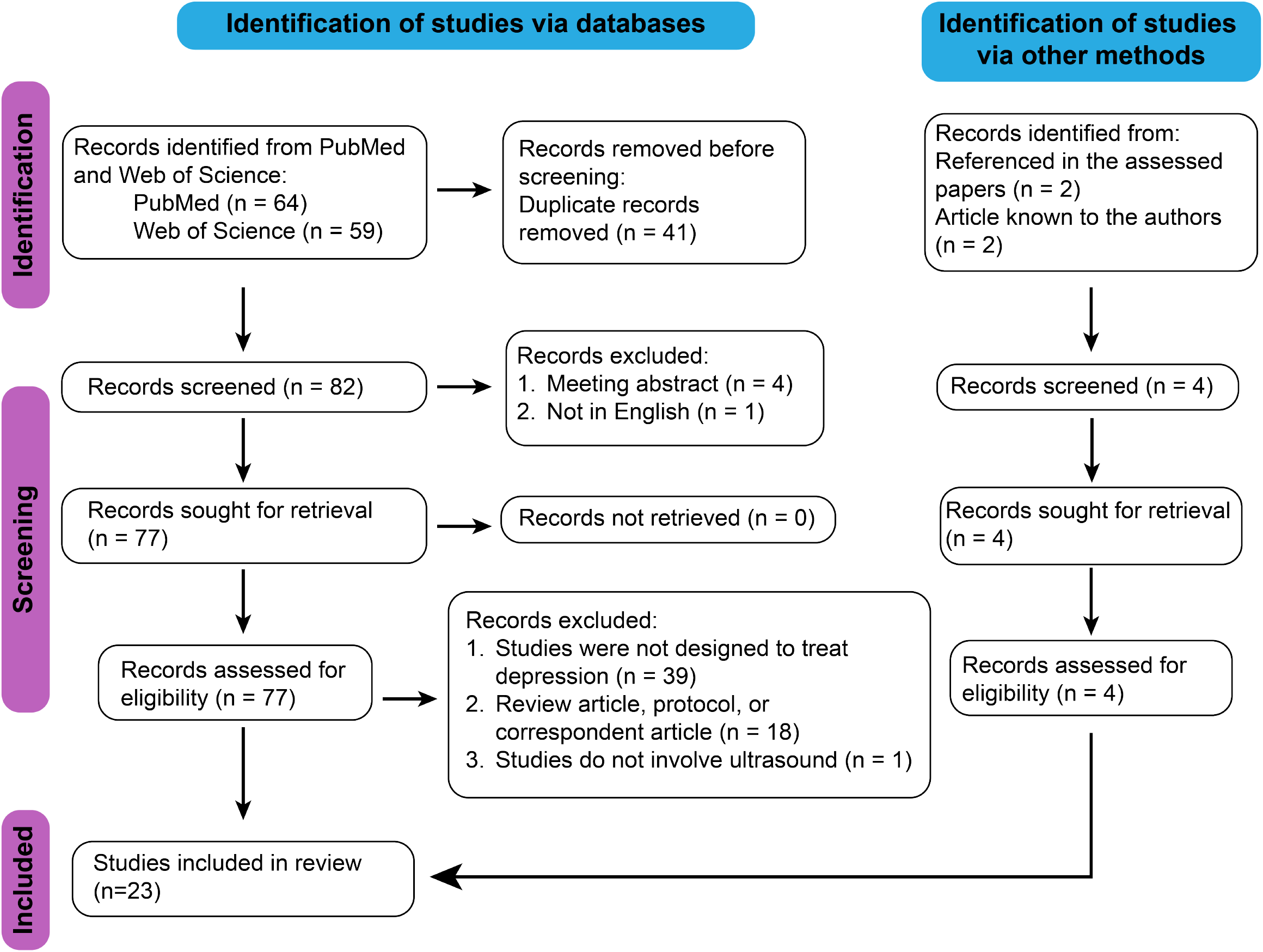
PRISMA flow diagram for study identification and selection.

### Search strategy

We conducted a systematic literature search using PubMed and Web of Science. The first search was conducted on December 10, 2024. We searched peer-reviewed papers that contain the keyword *Ultrasound* in the title AND the keywords *Ultrasound, neuromodulation, treatment*, and *depression* in all fields. Individual additional articles known to the authors were also added. We conducted a second literature search on June 20, 2025, during the manuscript revision, using the same databases and search strategy, except that we removed the requirement for the keyword *treatment* in all fields to capture potentially relevant studies.

### Eligibility criteria

We included studies that investigated the effects of ultrasound-based interventions in either (1) human subjects diagnosed with MDD or (2) depression-like animal models. Eligible study designs included randomized controlled trials, prospective cohort studies, and case-control studies published in full-text English. For human studies, eligibility required reporting of clinical or functional outcomes, such as changes in depression severity measured by scales (e.g., Hamilton Depression Rating Scale), neuroimaging findings, electrophysiological measures, or biochemical markers. For animal studies, eligibility required assessment of depression-like behaviors or neurobiological readouts such as gene expression or immunohistochemistry. Given the lack of consensus on optimal stimulation targets for LIFUS in depression, we did not restrict inclusion based on ultrasound targets. This decision is in line with a systematic review conducted during the early development of deep brain stimulation for depression to evaluate the general therapeutic potential^50^.

### Study selection

One reviewer removed duplicate reports (n = 41) before the study selection. Thus, a total of 86 articles (*n*=82 articles retrieved from databases and *n*=4 additional articles) were included in the study selection screen. We assessed the eligibility of each retrieved study in two phases: (1) a screen of title and abstract, and (2) a full-text review. The first reviewer sent the assessment of eligibility to the second reviewer, and any disagreements in the study selection were thoroughly discussed. The third reviewer made a final decision about study eligibility if the first and second reviewers did not reach consensus. Application of all selection criteria resulted in the removal of 63 articles, which left 23 articles that were subjected to data extraction and statistical analyses.

### Quality assessment

To ensure a rigorous evaluation of the included studies, we first assessed risk of bias and methodological quality using standardized tools appropriate to the study design. We applied RoB 2 for randomized controlled trials, ROBINS-E for open-label clinical studies, and SYRCLE’s risk of bias tool for preclinical animal studies^51–53^. Recognizing that these tools differ in structure, such as types of bias domain and signal questions, we developed a unified framework derived from the standardized tools (**Appendix 1**). This framework assessed each of the 23 included studies for risk of bias based on three categories: selection, comparability, and outcome. The maximum score for each category is 1. Good studies were defined as those that scored 1 in one category and at least 0.5 in the other two categories. Poor studies were defined as those that scored 0 in any one of the three categories. We consider the remaining studies of moderate quality. All reviewers discussed and resolved any disagreements on quality assessment.

### Data extraction

For human studies, we extracted the mean and variance of standard depression scales, including the Hamilton Depression Rating Scale and the Montgomery-Åsberg Depression Rating Scale, at baseline and the latest follow-ups. We also extracted the number of responders. For animal model studies, we extracted the mean and variance of measures for depressive-like behaviors, including immobility time in the forced swim test and the tail suspension test, and the sucrose preference index in the sucrose preference test. For all included studies, we extracted the sample size and the reported adverse effects. We extracted intent-to-treat (ITT) estimates and per-protocol estimates separately. Among the included LIFUS studies, only Riis et al. reported both ITT and per-protocol estimates^37^. In Oh et al., only per-protocol estimates were available because three subjects dropped out and no outcome data were collected for them^28^. The remaining human and animal studies did not report any subject exclusions or protocol deviations; therefore, we treated the available data from these studies as ITT estimates. Unless otherwise specified, the meta-analysis results presented in the Results section are based on ITT estimates. As a sensitivity analysis, we conducted a separate meta-analysis using per-protocol estimates and compared these results to the primary analysis to assess robustness. When multiple publications reported findings from the same dataset, we included only the most comprehensive study (e.g., the one with the largest sample size or most complete follow-up data).

### Statistical analysis

We retrieved or inferred the effect size (e.g., Cohen’s d) between the ultrasound treatment and control groups or between baseline and follow-up data. When available, we extracted means, standard errors (SE) or standard deviation (SD), and sample sizes to estimate the effect size *d* and its variance *Var*(*d*) (Eqs. 1-3). In studies that conducted Analysis of Variance (ANOVA) and reported F-statistics and degrees of freedom for the treatment effect *df*_*effect*_ and residual degrees of freedom *df*_*error*_, we estimated effect sizes and variances based on Eqs. 3-6^54^. For studies where means and variances were not reported but t-statistics were available, we estimated the effect size *d* and its variance *Var*(*d*) using Eqs. 3 and 7. For one study^40^ that only reported the change in depression scale from baseline to follow-up and its standard error, we estimated the effect size and its variance based on the formulas provided in^55^. Given heterogeneity across studies, we used random-effects models as the primary approach to estimate the pooled effect size. This approach incorporates both within-study and between-study variance. We used the DerSimonian-Laird estimator of between-study variance 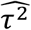 (Eq. 8). We estimated pooled effect size 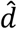 under the random-effects model using Eq. 9, where *k* denotes the number of studies. We estimated the 95% confidence interval (CI) as 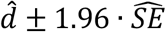, where 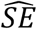 is the standard error of the pooled effect size, calculated according to Eq. 10. For reference, we included meta-analysis results from fixed-effects models in Appendix 2. For the fixed-effect model, we estimated 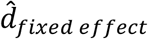 and 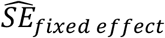 according to Eqs. 11 and 12, with inverse-variance weighting. We evaluated heterogeneity across studies by calculating the *I*^2^ metric^56^. A study of low-intensity pulsed ultrasound in a rat model of chronic stress tested three pulse frequencies (200, 285, and 500 Hz) and reported a significant effect only at 200 Hz^29^. Due to non-significant effects at 285 Hz or 500 Hz and the lack of exact statistics for these frequencies, we assumed their effect sizes to be zero and synthesized the results to estimate the overall effect before pooling data across studies. In human studies, we adhered to the definition of responder as a 50% reduction in the standard depression scale value^14^. We synthesized the number of responders and total subjects across studies to calculate the overall response rate. Given different stimulation targets used, we also synthesized the response rate for each targeted brain region. For human randomized trials that reported depression scale at baseline and after the treatment for both active and sham treatment groups, we estimated the effect size of ultrasound treatment and the effect size of ultrasound treatment compared with sham treatment, both adjusted for baseline. We defined the overall effect size as large for 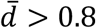, medium for 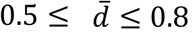, and small for 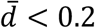.

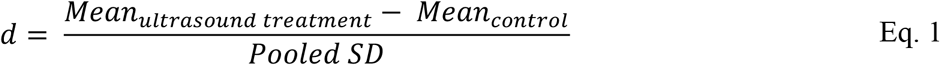

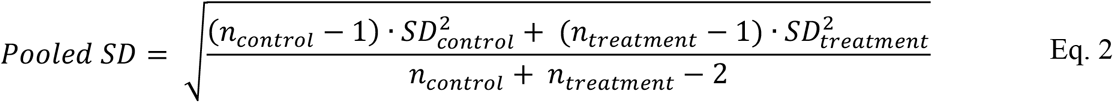

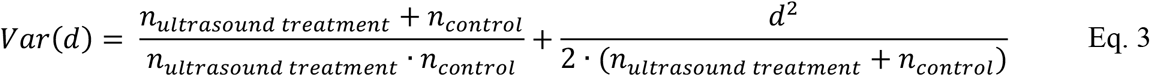

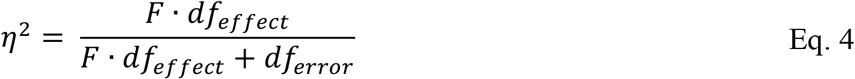

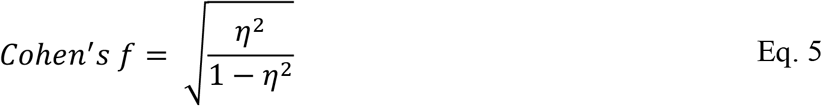

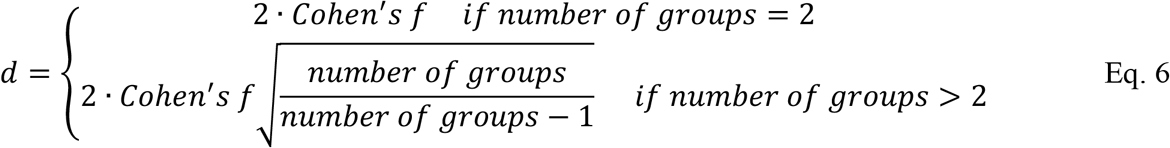

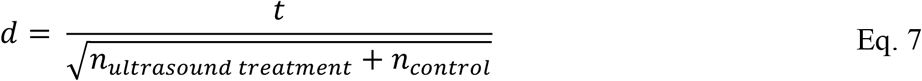

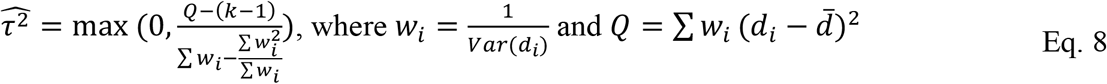

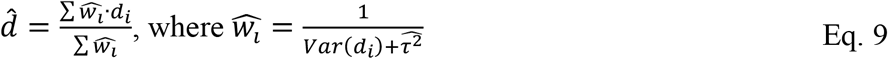

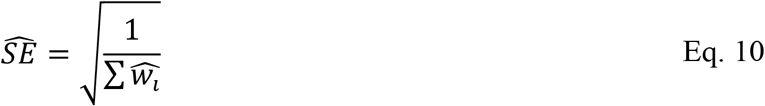

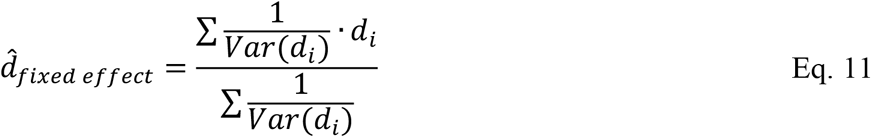

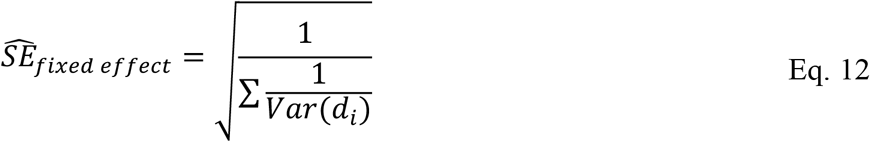

## Results

This review analyzed data from 23 peer-reviewed articles investigating ultrasound applications in the treatment of depression or depression-like behaviors (**Table 1**). The 23 peer-reviewed articles comprise 14 human studies and 9 studies using animal models. The 13 human studies report results from five trials, including six registered clinical trials (NCT02348411, NCT05301036, NCT04405791, NCT06085950, NCT06320028, and

**Table 1.** Summaries of study population, approach, outcomes, and efficacies reported in the included studies. We grouped studies into rodent LIFUS studies, human MRgFUS studies, and human LIFUS studies. We extracted spatial-peak pulse-average intensity (Isppa) when available; if Isppa was not reported, we extracted spatial-peak temporal-average intensity (Ispta). “NA” indicates data not available or not inferable. Unless otherwise specified, “control” in rodent models refers to animals that did not undergo procedures to induce depression-like behaviors, while “sham” refers to animals that received sham (inactive) stimulation.

| Author, year | Study population | (Stimulation) protocol | Outcome measures | Efficacy | Additional findings |
| --- | --- | --- | --- | --- | --- |
| **Preclinical studies (rodent models)** |  |  |  |  |  |
| Wu et al.<sup>42</sup>, 2023 | Repeated corticosterone-induced depression-like model: N = 8 (LIFUS), N = 8 (sham); vehicle-treated control: N = 8 | **Target:** dorsal raphe nucleus<br>**Intensity (Isppa):** 2.42 W/cm<sup>2</sup><br>**Fundamental frequency:** 1.1 MHz<br>**Pulse repetition frequency:** 1000 Hz<br>**Tone burst duration:** 0.5 ms<br>**Sonication duration:** 1 s<br>**Inter-stimulus interval:** 1 s<br>**Duration:** 30 minutes daily for two weeks | c-Fos immunoreactivity, sucrose preference index, immobility time in tail suspension test, time spent in elevated plus-maze test, and levels of serotonin. | Sucrose preference index was higher, and immobility time was reduced in LIFUS group compared to sham. | The H&E staining showed no damage or hemorrhage. The number of c-Fos-positive cells in the dorsal raphe nucleus was higher in the LIFUS group. |
| Hou et al.<sup>43</sup>, 2024 | Chronic restraint stress-induced depression-like model: N = 8 (control), N = 8 (saline), N = 8 (saline+ultrasound), N = 9 (PEGylated gas vesicles), and N = 9 (PEGylated gas vesicles+US) | **Target:** dorsal raphe nucleus<br>**Intensity (Isppa/Ispta):** NA (Peak pressure: 0.13-0.30 MPa)<br>**Fundamental frequency:** 1 MHz<br>**Pulse repetition frequency:** 1000 Hz<br>**Tone burst duration:** 0.3 ms<br>**Sonication duration:** 0.3 s<br>**Inter-stimulus interval:** 10 s<br>**Duration:** NA | Immobility time in the forced swim test and tail suspension test. | Mice receiving LIFUS exhibited reduced immobility time. | Ultrasound induced a 2.5-fold increased expression of c-Fos in 5-HT neurons compared to the control condition. |
| Yi et al.<sup>27</sup>, 2022 | Lipopolysaccharide-induced depression-like model: N = 13 (LIFUS), N = 12 (sham); control: N = 12 | **Target:** ventromedial prefrontal cortex<br>**Intensity (Isppa):** 10.09 W/cm<sup>2</sup><br>**Fundamental frequency:** 0.5 MHz<br>**Pulse repetition frequency:** 100 Hz<br>**Tone burst duration:** 5 ms<br>**Sonication duration:** 60 s<br>**Inter-stimulus interval:** 120 s<br>**Duration:** Single session of 30 minutes | c-Fos immunoreactivity in prefrontal neuron, immobility time in forced swimming test, and immobility time in tail suspension test. | LIFUS reduced immobility time compared to sham. | LIFUS did not cause any damage in prefrontal cortex based on H&E and Nissl staining. LIFUS increased the expression of c-Fos. |
| Zhu et al.<sup>30</sup>, 2023 | Chronic restraint stress-induced depression-like model: N = 11 (LIFUS), N = 11 (sham); control: N = 10 | **Target:** dorsal raphe nucleus<br>**Intensity (Isppa):** 5.68 W/cm<sup>2</sup><br>**Fundamental frequency:** 1.1 MHz<br>**Pulse repetition frequency:** 1000 Hz<br>**Tone burst duration:** 0.5 ms<br>**Sonication duration:** 1 s<br>**Inter-stimulus interval:** 1 s<br>**Duration:** 30 minutes daily for 14 days | c-Fos immunoreactivity, sucrose preference index, and immobility time in tail suspension test. | LIFUS reduced immobility time compared with the sham group. | H&E staining did not reveal tissue damage. c-Fos positive cells expression levels and the 5-HT levels in the dorsal raphe nucleus increased after LIFUS. |
| Zhang et al.<sup>44</sup>, 2018 | Chronic restraint stress-induced depression-like model: N = 17 (LIFUS), N = 16 (sham); control with sham LIFUS: N = 16 | **Target:** prelimbic cortex<br>**Intensity (Isppa):** 7.59 W/cm<sup>2</sup><br>**Fundamental frequency:** 0.5 MHz<br>**Pulse repetition frequency:** 1500 Hz<br>**Tone burst duration:** 0.4 ms<br>**Sonication duration:** 0.4 s<br>**Inter-stimulus interval:** 3 s<br>**Duration:** 15 minutes daily for 2 weeks | Sucrose preference index, immobility time in forced swimming test, distance travelled in open field test, brain-derived neurotrophic factor in left hippocampus. | LIFUS stimulation led to higher sucrose preference index, increased distance traveled, and reduced immobility time. | H&E staining confirmed no tissue damage or hemorrhage after sonication. LIFUS increased the expression of brain-derived neurotrophic factor. |
| Herlihy et al.<sup>58</sup>, 2023 | 6-hydroxydopamine (6-OHDA) lesion- | **Target:** celiac plexus (vagus nerve)<br>**Intensity (Isppa):** NA | Immobility time in forced swim test and sucrose preference index. | LIFUS increased sucrose preference but had no significant | NA |
|  | based depression-like model. Animals were divided into four conditions: control, control+LIFUS, depression-like model, and depression-like model+LIFUS. N = 10-11/per conditions for the forced swim test. N = 5, 13, 18, 22 for the sucrose preference test. | **Fundamental frequency:** 2.5 MHz<br>**Pulse repetition frequency:** 5 Hz<br>**Tone burst duration:** 0.12 ms<br>**Sonication duration:** NA<br>**Inter-stimulus interval:** NA<br>**Duration:** 3 minutes, 6 h apart, over 5 consecutive days |  | effect on immobility time. |  |
| Wang et al.<sup>41</sup>, 2024 | Chronic restraint stress-induced depression-like model: N = 16 (LIFUS), N = 14 (sham); control with sham LIFUS: N = 14 | **Target:** ventral tegmental area<br>**Intensity (Ispta):** 150 mW/cm<sup>2</sup><br>**Fundamental frequency:** 0.5 MHz<br>**Pulse repetition frequency:** 1500 Hz<br>**Tone burst duration:** 0.3 ms<br>**Sonication duration:** 0.2 s<br>**Inter-stimulus interval:** 1.6 s<br>**Duration:** 15 minutes daily for 10 days | Immobility time in forced swimming test, Immobility time in tail suspension test, sucrose preference index in sucrose preference test, dopaminergic release in prefrontal cortex based on fiber photometry, and the number of dopaminergic neuron in ventral tegmental area. | LIFUS improved all measured depression-like behavior and increased dopamine levels in prefrontal cortex. | NA |
| Zhang et al.<sup>29</sup>, 2021 | Chronic unpredictable stress model: N = 8 (LIFUS), N = 8 (sham) | **Target:** ventromedial prefrontal cortex<br>**Intensity (Isppa):** 3.84 W/cm<sup>2</sup><br>**Fundamental frequency:** 0.8 MHz<br>**Pulse repetition frequency:** 200 Hz, 285 Hz, and 500 Hz<br>**Tone burst duration:** 0.2 ms<br>**Sonication duration:** 1 s<br>**Inter-stimulus interval:** 3 s<br>**Duration:** 20 minutes daily for four weeks | Immobility time in the forced swimming test, sucrose preference index, distance traveled in open field test, and c-Fos immunoreactivity. | Only LIFUS with 200 Hz pulse repetition frequency increased sucrose preference index, reduced forced swim immobility time in stressed rats. LIFUS had no significant effect on distance traveled in open field test. | The number of c-Fos-positive cells in the ventromedial prefrontal cortex was increased. Histological staining revealed no gross tissue damage. |
| Ren et al.<sup>59</sup> | Same as Zhang et al.<sup>29</sup>, 2021 | Same as Zhang et al.<sup>29</sup>, 2021 | Transcriptome profile. | Depression-like behavior assessment was reported in Zhang et al.<sup>29</sup>, 2021 | LIFUS reversed the abnormal expression of genes linked to glucose and lipid metabolism in the hippocampus and prefrontal cortex in model rats. |
| **Human studies (MRgFUS)** |  |  |  |  |  |
| Kim et al.<sup>35</sup>, 2018 | N = 1 (MDD patient) | MRgFUS for anterior capsulotomy using the ExAblate 4000 device (InSightec, Haifa, Israel).<br>Intensity (Isppa/Ispta): NA | Hamilton Depression Rating Scale score and Beck Depression Inventory score. | MRgFUS improved depression scale scores. | NA |
| Hamani et al.<sup>36</sup>, 2024 | N = 35 were offered MRgFUS: N = 12 (MDD patients), N = 15 (patients diagnosed with obsessive compulsive disorder), N = 8 (insufficient heating to create a lesion) | MRgFUS for anterior capsulotomy using the ExAblate 4000 device (InSightec, Haifa, Israel).<br>Intensity (Isppa/Ispta): NA (targeting temperature > 53°C) | Clinical evaluation for adverse effects, Hamilton Depression Rating Scale scores. | Non-significant reduction in Hamilton Depression Rating Scale scores compared to baseline in MDD patients at 6 and 12 months. | No serious adverse effects were reported. Nonserious adverse events occurred in 7 patients, including transient headaches and pin-site swelling. |
| Davidson et al.<sup>60</sup>, 2021 | A subset of patients included in Hamani et al.<sup>36</sup>, 2024 | Same as Hamani et al.<sup>36</sup>, 2024 | Treatment success ratio defined by visible bilateral lesions on postoperative MRI. | Complete clinical outcomes was reported in Hamani et al.<sup>36</sup>, 2024. | The treatment success ratio was 15/22. |
| Davidson et al.<sup>61</sup>, 2020 | A subset of patients included in Hamani et al.<sup>36</sup>, 2024 | Same as Hamani et al.<sup>36</sup>, 2024 | Procedural considerations involved in performing bilateral MRgFUS capsulotomy. | Complete clinical outcomes was reported in Hamani et al.<sup>36</sup>, 2024. | Greater acoustic energy is needed for treatment of the second side, possibly due to skull heating. |
| Davidson et al.<sup>62</sup>, 2020 | A subset of patients included in Hamani et al.<sup>36</sup>, 2024 | Same as Hamani et al.<sup>36</sup>, 2024 | Cognitive performance based on neuropsychological tests. | Complete clinical outcomes was reported in Hamani et al.<sup>36</sup>, 2024. | Postoperative cognitive scores were stable or improved at both 6 and 12 months postoperatively compared to baseline. |
| Davidson et al.<sup>14</sup>, 2020 | A subset of patients included in Hamani et al.<sup>36</sup>, 2024 | Same as Hamani et al.<sup>36</sup>, 2024 | Clinical outcomes, cognitive testing, and structural imaging. | Complete clinical outcomes was reported in Hamani et al.<sup>36</sup>, 2024. | NA |
| **Human studies (LIFUS)** |  |  |  |  |  |
| Oh et al.<sup>28</sup>, 2024 | N = 26<br>randomized: N = 11 (LIFUS), N = 12 (sham), N = 3 (dropout) | **Target:** dorsal lateral prefrontal cortex<br>**Intensity (Isppa):** 600 mW/cm<sup>2</sup><br>**Fundamental frequency:** 0.25 MHz<br>**Pulse repetition frequency:** 500 Hz<br>**Tone burst duration:** 1 ms<br>**Sonication duration:** 0.3 s<br>**Inter-stimulus interval:** 5.7 s<br>**Duration:** 15 minutes daily for 10 days<br>**Duration:** 20-minute session, twice weekly for two weeks | Montgomery-Åsberg Depression Rating Scale and Resting-state functional magnetic resonance imaging. | LIFUS reduced Montgomery-Åsberg Depression Rating Scale and increased connectivity between subgenual anterior cingulate cortex and prefrontal cortex. | NA |
| Riis et al.<sup>37</sup>, 2024 | N = 22<br>randomized: N = 9 (LIFUS), N = 12 (sham), N = 3 (deviation from protocol) | **Target:** subcallosal cingulate<br>**Intensity (Isppa):** 31.1 W/cm<sup>2</sup><br>**Fundamental frequency:** 0.65 MHz<br>**Pulse repetition frequency:** 100 Hz<br>**Tone burst duration:** 5 ms<br>**Sonication duration:** 0.03 s<br>**Inter-stimulus interval:** 1.4 s or 0.7 s<br>**Duration:** 3 blocks (A, B, C). Block A contained three to five 1-minute LIFUS to test the tolerability. Block B contained six 3-minute LIFUS with 1.4 s inter-stimulus intervals. Block C contained six 3-minute LIFUS with 0.7 s inter-stimulus intervals. | Sadness subscale of the expanded Positive and Negative Affect Schedule (PANAS-X), Hamilton Depression Rating Scale score-6 (HDRS-6), blood oxygenation level-dependent (BOLD) signal. | In the per-protocol sample, LIFUS group had greater reduction in the PANAS-X score immediately following treatment, and greater reduction in HDRS-6 scores at 24s hour, but not at 7 days. | Functional MRI shows deactivation in the subcallosal cingulate cortex, and activation in the left ventrolateral prefrontal cortex and bilateral temporal cortex. No severe adverse events occurred during or immediately after stimulation. Two subjects in the LIFUS group experienced a severe psychiatric adverse event 24-hour after treatment. |
| Riis et al.<sup>63</sup>, 2023 | N = 1 (treatment-resistant depression patient) | **Target:** subcallosal cingulate<br>**Intensity (Isppa/Ispta):** NA (Estimated peak pressure: 1 MPa)<br>**Fundamental frequency:** 0.65 MHz<br>**Pulse repetition frequency:** NA<br>**Tone burst duration:** NA<br>**Sonication duration:** 30 ms<br>**Inter-stimulus interval:** 4 s<br>**Duration:** single session of 64 minutes | Hamilton Depression Rating Scale score. | LIFUS decreased Hamilton Depression Rating Scale score, which remained low for 44 days following the sonication. | No adverse effects were reported. |
| Riis et al.<sup>57</sup>, 2024 | N = 2 (treatment-resistant) | **Target:** subcallosal cingulate<br>**Intensity (Isppa):** 31.1 W/cm<sup>2</sup> | Non-standard Depression and | LIFUS improved depression and anxiety. | No adverse effect or apparent changes in |
|  | depression patients) | **Fundamental frequency:** 0.65 MHz<br>**Pulse repetition frequency:** NA<br>**Tone burst duration:** NA<br>**Sonication duration:** 30 ms<br>**Inter-stimulus interval:** 4 s<br>**Duration:** single session of 60 – 180 s | anxiety rating assessed. |  | structural imaging were reported. Targeting accuracy was on a scale of millimeter. |
| Li et al.<sup>34</sup>, 2020 | N =30 (sham, LIFUS+TMS), N = 30 (LIFUS) | **Targets:** lateral orbitofrontal cortex, dorsal prefrontal cortex, and cuneiform lobe.<br>**Intensity (Ispta):** 720 mW/cm<sup>2</sup><br>**Fundamental frequency:** 0.65 MHz<br>**Pulse repetition frequency:** 100 Hz,<br>**Tone burst duration:** 0.5 ms<br>**Sonication duration:** 30 s<br>**Inter-stimulus interval:** 30 s<br>**Duration:** The sham group underwent 30 minutes TMS treatment, The LIFUS group underwent 15 minutes TMS, followed by 15 minutes LIFUS for each target, 5 days per week, for 2 months. | Hamilton Depression Rating Scale score. | Hamilton Depression Rating Scale score was lower in the treatment group than in the sham group. | No statistical difference in adverse reactions between the treatment and sham groups. |
| Attali et al.<sup>38</sup>, 2025 | N = 5 (MDD patients) | **Target:** subcallosal cingulate white matter tracts<br>**Intensity (Ispta):** 184.4 mW/cm<sup>2</sup><br>**Fundamental frequency:** 0.5 MHz<br>**Pulse repetition frequency:** 14 Hz<br>**Tone burst duration:** 4.5 ms<br>**Sonication duration:** 5 s<br>**Inter-stimulus interval:** 15 s<br>**Duration:** 5-minute LIFUS, 5 times per day for 5 days. | Montgomery-Åsberg Depression Rating Scale (MADRS) and resting state functional connectivity. | Significant reduction in MADRS at the end of the treatment week compared to the baseline. | No serious adverse events occurred. No sensation or pain were reported. Increased functional connectivity between subcallosal cingulate and left dorsolateral prefrontal cortex and decreased connectivity between subcallosal cingulate and the right hippocampal region. |
| Schachter et al.<sup>40</sup>, 2025 | N = 20 diagnosed with moderate to severe depression) | **Target:** medial prefrontal cortex<br>**Intensity (Ispta):** 670 mW/cm<sup>2</sup><br>**Fundamental frequency:** 0.4 MHz | Beck Depression Inventory-II (BDI-II) , Perseverative Thinking Questionnaire | BDI-II and HDRS rating reduced after LIFUS intervention. | No observed adverse event. The mean subjective report of adverse sensation (pain |
|  |  | **Pulse repetition frequency:** 10 Hz,<br>**Tone burst duration:** 5 ms<br>**Sonication duration:** 5 s<br>**Inter-stimulus interval:** 15 s<br>**Duration:** 10-minute session, five sessions during the first week, followed by three sessions per week for the subsequent two weeks | (PTQ), and Hamilton Depression Rating Scale (HDRS). |  | and tension) is about 2 out of 10. |
| Cheung et al.<sup>39</sup>, 2025 | N = 30 randomized: N = 15 (LIFUS), N = 15 (waitlist control) | **Target:** dorsolateral prefrontal cortex<br>**System:** NEUROLITH, Storz Medical AG, Tägerwil, Switzerland<br>**Intensity (Ispta):** NA (Energy level: 0.2–0.25 mJ/mm<sup>2</sup>)<br>**Fundamental frequency:** 0.33 MHz<br>**Pulse repetition frequency:** 4–5 Hz<br>**Tone burst duration:** single ultrasound pulse (3 μs)<br>**Sonication duration:** 300 pulses<br>**Inter-stimulus interval:** NA<br>**Duration:** six 30-minute sessions with three sessions per week on alternate days for two weeks | Hamilton depression rating scale (HDRS)-17. | HDRS rating was significantly different between treatment and control groups after intervention. | Sustained treatment effect was observed at the 3-month follow-up in all subjects who received LIFUS intervention. |

NCT03421574, NCT05006365). The 9 animal studies used mice and rats, and five of these used restraint stress models. The rodent models in the remaining four studies include chronic corticosterone-induced depression-like models, chronic unpredictable stress depression-like models, Lipopolysaccharide-induced depression-like models, and 6-hydroxydopamine (6-OHDA) lesion-induced anhedonia-like behaviors. For the latest reports of four independent randomized controlled trials investigating LIFUS neuromodulation, the ROB2 assessment indicated a low risk of bias or some concerns (see **source data – ROB2 assessment** for details). ROBINS-E assessment for open-label human studies indicated a high risk of bias in one study and some concerns in others. The primary sources of bias were selection bias and bias arising from outcome measurement, as participants and investigators were aware of the intervention (see **source data – ROBINS-E Tool** for details). For preclinical studies, SYRCLE’s risk of bias tool identified that most animal studies had risk of bias related to group allocation and blinding, as many did not explicitly state whether allocation was concealed from the investigators or whether outcome assessors were blinded (see **source data – animal SYRCLE’s risk of bias** for details). The unified quality evaluation of the 23 studies indicated that 14 are of good quality, 6 are of moderate quality, and 3 are of poor quality (**Supplementary Figure 1**)

### HIFUS thermal ablation to treat depression

Both LIFUS and HIFUS have been used to treat human patients with major depressive disorders (**Figure 2A**). HIFUS was applied to ablate the anterior limb of the internal capsule in patients with MDD. This technique, called magnetic resonance-guided focused ultrasound (MRgFUS), directs ultrasound to generate heat at the target site and cause coagulative necrosis and tissue ablation (**Figure 2B**). MRgFUS creates intracranial lesions without requiring a cranial window through craniotomy. The US Food and Drug Administration (FDA) approved MRgFUS for the treatment of refractory essential tremor in July 2016. Subsequently, MRgFUS has been investigated as a non-invasive ablation method for other neurological disorders, including MDD, Obsessive-compulsive disorder, and brain tumors^64^. In human studies included in this review, eligible patients for MRgFUS are those with treatment-resistant MDD. These studies utilized MRgFUS to target the anterior limb of the internal capsule, a white matter region that contains fibers from the prefrontal cortex towards the ventral striatum and the thalamus. The therapeutic mechanism mediating MRgFUS capsulotomy involves the disruption of the output from the anterior nucleus of the thalamus projecting to the paraterminal gyrus tract while preserving the dorsolateral prefrontal-thalamic tracts to avoid causing frontal lobe syndrome. The MRgFUS procedure is administered as follows. Patients are positioned in an intraoperative MRI scanner with a stereotactic frame. Low-energy sonication pulses are delivered to induce temperatures of 40°–42°C, which serves to verify targeting accuracy before applying HIFUS. Temperature feedback from MR thermometry allows the neurosurgical team to make millimeter-scale adjustments to ensure that the focal point of heating aligns with the intended target region. Following successful verification, high-power sonication pulses are applied iteratively, raising temperatures to between 50–56°C for durations exceeding three seconds.

**Figure 2.**
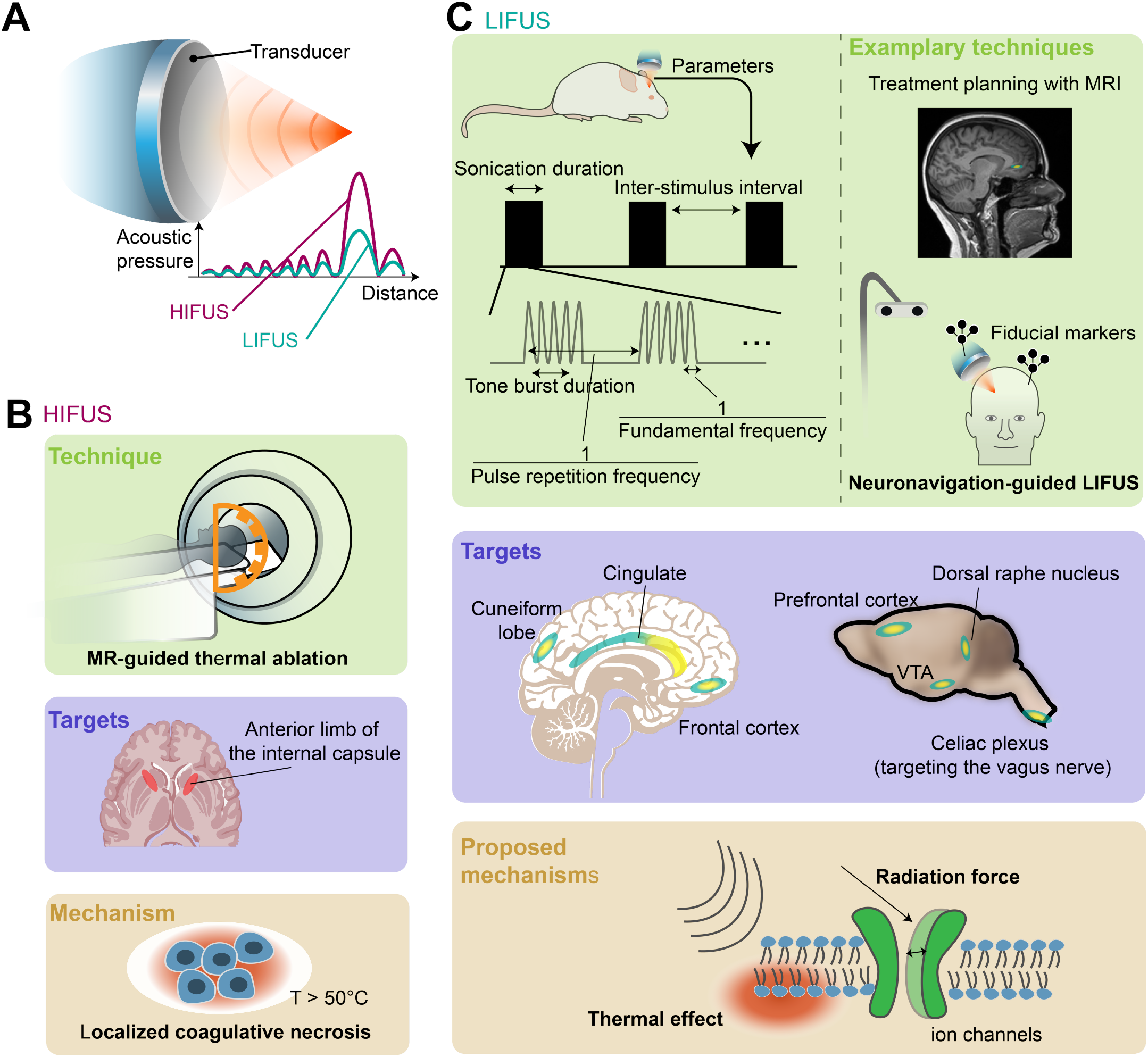
Summary of techniques, targets, and potential mechanisms in the application of high-intensity focused ultrasound (HIFUS) thermal ablation and low-intensity focused ultrasound (LIFUS) neuromodulation for treating depression. **A.** Ultrasonic applications can be categorized into LIFUS or HIFUS based on the intensity (e.g., acoustic pressure). Each category involves distinct techniques and mechanisms. **B**. HIFUS was primarily used for thermal ablation in humans. MR: magnetic resonance. **C**. LIFUS neuromodulation involves five independent parameters: fundamental frequency, pulse repetition frequency, tone burst duration, inter-stimulus interval, and sonication duration. Generally, administering LIFUS requires neuroimaging, acoustic simulation, and an optical tracking-based neuronavigation system^28,38–40^. A slightly different protocol involves mechanically registering the device to the subject’s head and calibrating the beamforming based on the first MR scan^57^. VTA: ventral tegmental area. The primary mechanisms mediating LIFUS are thought to be radiation force (membrane deformation and mechanosensitive ion channel activation caused by ultrasound) and thermal effects.

### LIFUS neuromodulation to treat depression

In contrast to HIFUS, LIFUS aims to modulate neuronal activity without creating irreversible lesions. Its neuromodulation effects are thought to be transient. Most neuroimaging and physiological studies reported post-LIFUS effects lasting from 10 minutes to 2 hours^26,65,66^. The durability of its neuromodulation effect requires further research, as it may depend on the stimulation parameters. MDD is associated with dysregulation of neurotransmitter systems and neural circuits involved in mood regulation, stress response, and reward processing. LIFUS is gaining attention as a non-invasive neuromodulation technique with high spatial resolution and deep penetration depth for treating patients with drug-resistant MDD. Preclinical studies with depressive rodent models administer LIFUS via transcranial low-intensity pulsed ultrasound stimulation, which reduces the risk of thermal damage at the targeted brain region compared to continuous wave ultrasound. In these studies, the targeted brain regions include the dorsal raphe nucleus, prefrontal cortex, and ventral tegmental area. Three out of nine animal studies included in this review targeted the dorsal raphe nucleus to facilitate serotonin release^30,42,43^, while another four targeted the prefrontal cortex for its role in emotion regulation^27,29,44,59^. One study directed LIFUS toward the ventral tegmental area to regulate the dopamine system^41^. Beyond central targets, one study stimulated the vagus nerve by targeting the celiac plexus to harness the antidepressant effects of vagus nerve stimulation^58^. One out of the nine included rodent studies uniquely combined transcranial low-intensity ultrasound with PEGylated gas vesicles.

LIFUS neuromodulation protocol involves five independent parameters (**Figure 2C**): fundamental frequency, pulse repetition frequency, tone burst duration, inter-stimulus interval, and sonication duration. The fundamental frequency determines the oscillation rate of the ultrasound waves, which influences penetration depth. Pulse repetition frequency is the rate at which ultrasound pulses are emitted. The tone burst duration is the length of each ultrasound pulse, and the inter-stimulus interval is the time between consecutive pulses. The duty cycle represents the proportion of time the transducer actively emits ultrasound. It is determined by both the tone burst duration and pulse repetition frequency. A higher duty cycle may increase the risk of tissue heating. Transcranial low-intensity pulsed ultrasound has been applied in animal studies using a broad range of parameters: fundamental frequency ranging from 0.5–2.5 MHz, pulse repetition frequency ranging from 5–1.5 kHz, tone burst duration ranging from 0.2–5 ms, sonication duration ranging from 0.2–120 s, and inter-stimulus interval ranging from 1–120 s (**Table 1**).

Human trials are beginning to explore the use of transcranial low-intensity pulsed ultrasound stimulation to treat patients with MDD^28,34,38–40,57,63^. To accurately target brain regions based on individual neuroanatomy, clinical LIFUS protocols typically require neuroimaging, acoustic simulation, and an optical tracking-based neuronavigation system. Neuroimaging data, including Magnetic Resonance Imaging (MRI) and Computed Tomography (CT) scans, are acquired to define anatomical targets and simulate ultrasound propagation. Optionally, diffusion MRI can be used to identify patient-specific white matter targets^38^. Based on the skull properties derived from the CT scans, numerical simulations estimate the speed of sound, acoustic impedance, and the acoustic pressure field in the brain. These parameters can be used to define the transducer placement and compensate skull-induced aberrations of the ultrasound wave through changes in beamforming parameters or a personalized acoustic lens^38,57^. After treatment planning, the neuronavigation system registers the subject’s imaging space to physical space. This is typically achieved using a tracked stylus to localize anatomical landmarks, such as the nasion, or MRI-visible fiducials^28^ in the physical space. An optical marker is affixed to the LIFUS transducer, allowing the neuronavigation system to track its position and orientation in real-time. Additional markers are usually affixed to participants for continuous monitoring of the head position^39^. During the LIFUS session, the neuronavigation system provides feedback on the spatial offset between the transducer’s focal point and the planned target. In contrast to the neuronavigation approach, Riis et al. used a mechanical registration strategy. In their protocol, the patient’s head was immobilized with thermoplastic masks and mechanically coregistered with ultrasound transducers. An MRI is acquired, which includes both the subject’s brain anatomy and the transducer arrays with fiducial markers. They adjust the beamforming parameters to steer the ultrasound to the targeted brain area. In the subsequent treatment sessions, the subject’s head is placed in the same thermoplastic mask, and the transducer arrays are locked in the same mechanical positions. LIFUS targets include the subcallosal cingulate cortex and the dorsolateral prefrontal cortex. One study targeted the cuneiform lobe and the orbitofrontal cortex in addition to the prefrontal cortex^34^. The treatment protocol varied from study to study, with fundamental frequency ranges from 250–650 kHz and total intervention duration lasting from single sessions to multi-week regimens (**Table 1**). The LIFUS protocols reported in the included human trials in this review administered low-intensity pulsed stimulation without the use of microbubbles. These protocols are expected to have a low likelihood of inducing cavitation effects; therefore, we do not highlight cavitation effects in **Figure 2C**.

### Efficacy of HIFUS thermal ablation for treating depression

Our analysis of six studies in humans reporting the results of three clinical trials (NCT02348411, NCT03421574, and NCT03156335) concluded that MRgFUS successfully created a lesion (postoperative lesion >1 mm on MRI) for anterior capsulotomy in 28 of 36 subjects^35,36^. These subjects include patients with MDD or obsessive-compulsive disorder who underwent identical MRgFUS procedures. Unsuccessful lesioning primarily resulted from insufficient temperature elevation within the anterior limb of the internal capsule, under practical and safety considerations such as treatment duration and scalp heating. Key factors affecting the maximum temperatures achieved included skull density, skull thickness, and the angle of incidence^60^. A lower skull density and increased skull thickness were associated with lower achieved temperature. MRgFUS successfully created lesions in 13 patients with MDD. The clinical outcome data were available only for these 13 subjects. The following results regarding the efficacy of MRgFUS anterior capsulotomy are based on the per-protocol estimates. 5 of 13 patients met responder criteria (≥50% improvement from baseline on the Hamilton Depression Rating Scale) at 12 months postoperative, and 7 of 13 met responder criteria at 24 months at long-term (follow-up for up to 24 months postoperative). The reductions in Hamilton Depression Rating Scale is 28.13% ± 11.24% at 6 months postoperatively (compared with baseline, p = 0.027, t = 2.520, n = 13, paired t-test), and 22.98% ± 12.65% at 12 months postoperatively (compared with baseline, p = 0.071, t = 1.986, n = 13, paired t-test), and 39.37% ± 19.38 at long-term (compared with baseline, p = 0.093, t = 2.071, n = 6, paired t-test). No serious adverse events were reported for all patients undergoing MRgFUS for capsulotomy. Nonserious adverse events included transient headaches lasting for a few hours after MRgFUS, pin-site swelling, and a sensation of fogginess.

### Efficacy of LIFUS neuromodulation for treating depression

The nine rodent model studies in our review utilized three metrics to assess depressive-like behavior: immobility time in the forced swimming test (reported in 6 of 9 rodent studies), sucrose preference index (reported in 5 of 9 rodent studies), and immobility time in the tail suspension test (reported in 5 of 9 rodent studies). Two studies reported results from the same dataset, one focusing on depressive-like behavior and the other on transcriptomic profiles. We included only the study reporting the effects of LIFUS on depressive-like behavior in the meta-analysis. LIFUS had a large effect (Cohen’s d) on reducing immobility time in the forced swimming test (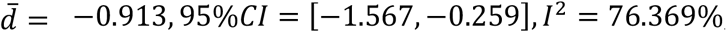, **Figure 3A**). The pooled effect size for the sucrose preference index was large (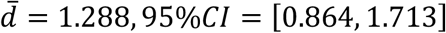, **Figure 3B**). However, there was substantial variability in effect sizes across studies (*I*^2^ = 62.461%). In addition, LIFUS reduced immobility time in the tail suspension test with a large effect (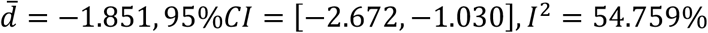, **Figure 3C**). Two rodent studies were based on the same dataset. Among the eight independent studies, five performed H&E staining and reported that no damage or hemorrhage was observed in the targeted tissue.

**Figure 3.**
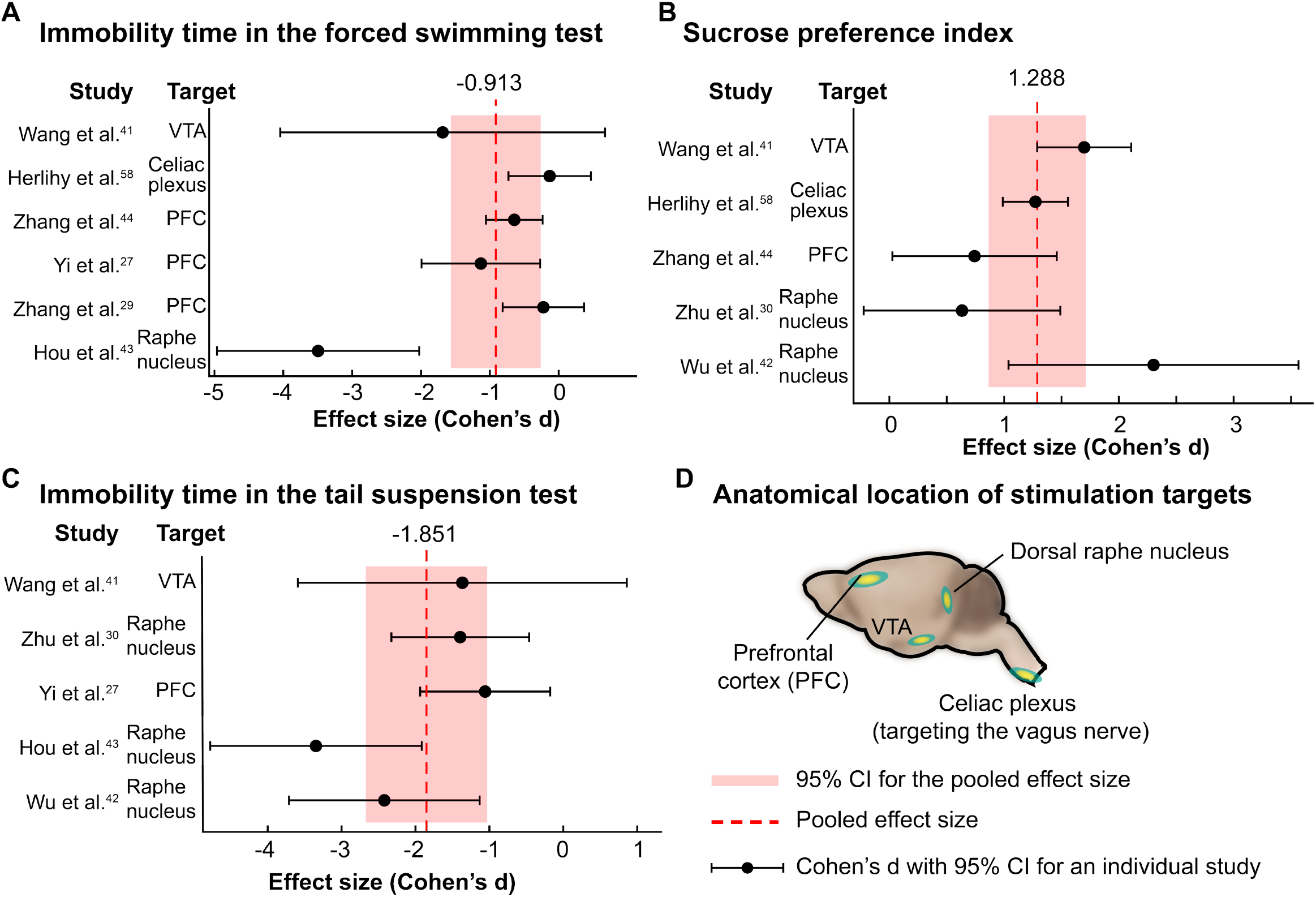
Efficacy of low-intensity focused ultrasound (LIFUS) stimulation in preclinical studies with rodent models. **A-C.** The effect of LIFUS neuromodulation on (**A**) immobility time in forced swimming test (seconds), (**B**) sucrose preference index (a.u.), and (**C**) immobility time in tail suspension test (s). The effect size was estimated using data before and after LIFUS stimulation. In Herlihy et al.^58^, the immobility time in the forced swimming test was not increased in the depressive rat model compared to healthy rats. **D**. Anatomical location of stimulation targets. VTA stands for ventral tegmental area.

This review evaluated 14 studies in humans, and eight of those investigated the effects of LIFUS neuromodulation in patients with MDD. One case study reported that LIFUS neuromodulation targeting the subcallosal cingulate cortex resolved depressive symptoms. The patient’s Hamilton Depression Rating Scale-6 score decreased from 11 to 0, and the patient remained in remission during 44 days of monitoring. One study reported that ultrasonic stimulation of the subcallosal cingulate cortex immediately reduced depression severity in two patients, as assessed by a psychiatrist. Two studies reported single-arm, open-label, non-randomized trials. The other four studies reported the results of randomized clinical trials and reported standard depression assessment at baseline and follow-ups for both active stimulation and sham groups. The synthesized response rate is 69.2% (54/78) in the treatment group, compared to 44.2% (23/52) in the sham group (Odds ratio = 2.837). In Li et al.^34^, the sham control group received 30-minute TMS treatment sessions, while the treatment group received sessions consisting of 15-minute TMS treatment and 15-minute LIFUS treatment. The synthesized response rate after excluding data from Li. et al. was 56.3% in the LIFUS treatment group, and 18.2% in the sham control group. LIFUS had a large effect on reducing depression severity within the ultrasound treatment group compared to the baseline depression score (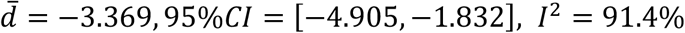, **Figure 4A**). LIFUS also had a large effect on reducing depression severity in the ultrasound treatment group when compared with the sham control group after adjusting for baseline scores (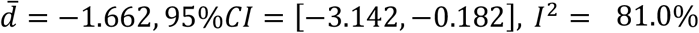 **Figure 4B**). To better understand the durability of LIFUS effects, we examined the timing of outcome assessments in the included study. Li et al., Cheung et al., and Schachtner et al. assessed outcomes only immediately after treatment, reporting a significant reduction in the depression rating scale compared to baseline or sham controls^34,39,40^. Oh et al. conducted a two-week follow-up and observed sustained effects^28^. Riis et al. conducted a one-week follow-up and found only a significant reduction in depression rating scale immediately after treatment, but not at follow-up^37^. Attali et al. reported the longest follow-up period (five weeks), observing peak effects immediately after treatment a gradual return to baseline by week five^38^. These findings suggest that LIFUS produces transient clinical effects, with duration varying by protocol.

**Figure 4.**
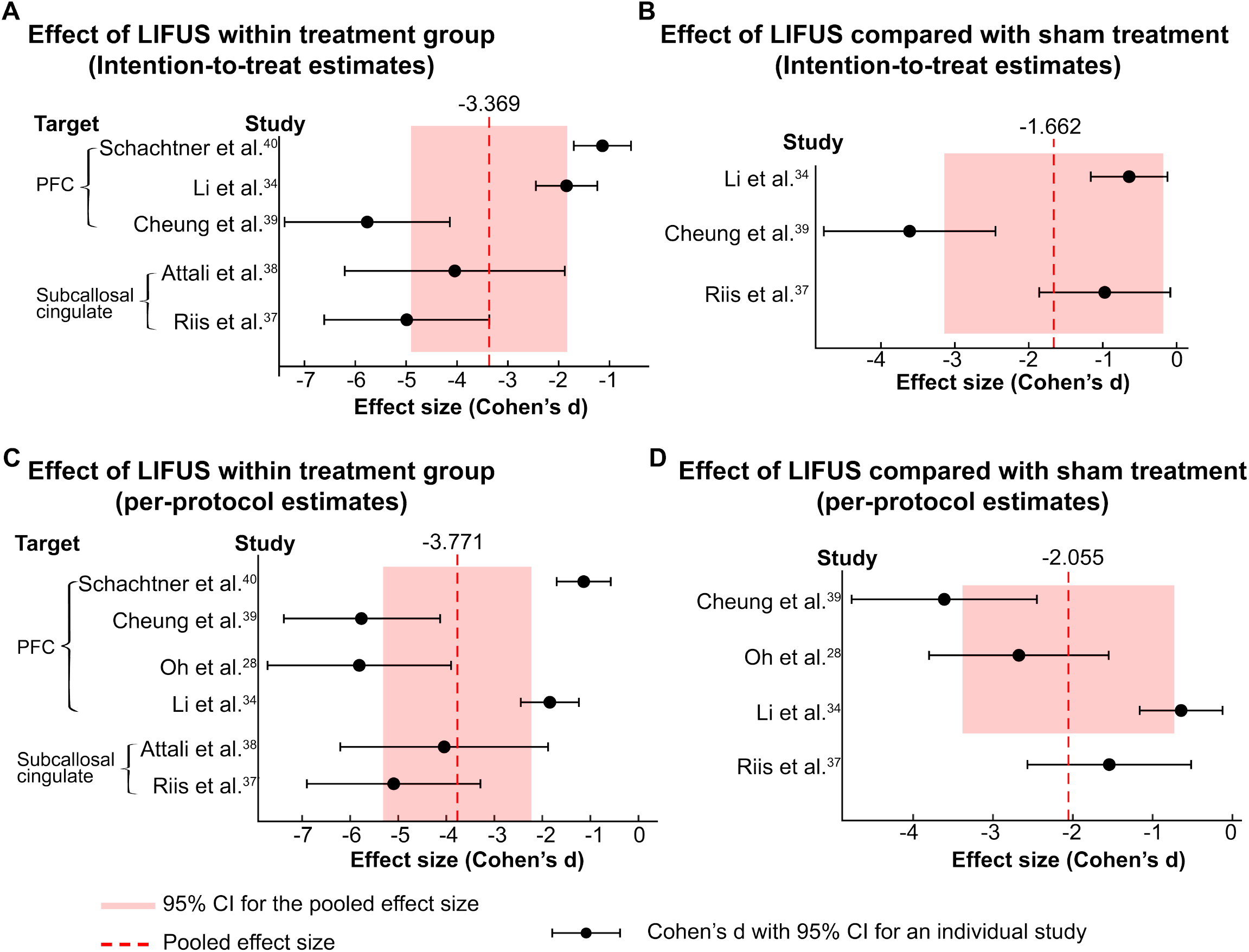
Efficacy of low-intensity focused ultrasound (LIFUS) neuromodulation in clinical trials. **A.** The effect of LIFUS on depression severity within the treatment group was assessed using intention-to-treat (ITT) depression scale scores before and after LIFUS treatment. In Li et al.^34^, LIFUS neuromodulation targeted the orbitofrontal, dorsal prefrontal cortex (PFC), and cuneiform lobe. Riis et al.^37^ and Attali et al.^38^ targeted the subcallosal cingulate cortex. Oh et al.^28^ and Cheung et al.^39^ targeted the dorsolateral PFC. Schachtner et al.^40^ targeted the anterior medial prefrontal cortex, which is anatomically close to the subcallosal cingulate cortex. **B**. Effect of LIFUS on the depression severity compared to sham treatment. The effect size represents the difference in changes in ITT depression scale scores for the LIFUS treatment and sham control groups. **C**. Effect of LIFUS on depression severity within the treatment group based on per-protocol depression scale scores before and after LIFUS treatment. **D**. Effect of LIFUS on the depression severity compared to sham treatment using per-protocol estimates.

A summary of findings using the GRADE approach to assess the certainty of the evidence is provided in **Table 2**. As a sensitivity analysis, we repeated the meta-analysis using per-protocol estimates. Similar to meta-analysis using ITT estimates, we found that LIFUS had a large effect within the ultrasound treatment group (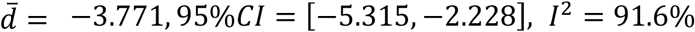, **Figure 4C**) and a large between-group effect (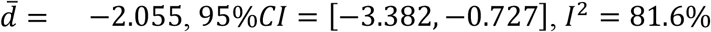, **Figure 4D**). LIFUS neuromodulation was well tolerated in the three randomized clinical trials, with 3/53 patients in the active treatment group and 4/52 patients in the sham control group reporting dizziness, vomiting, or diarrhea^28,34,37^. Two patients experienced severe psychiatric adverse events in the LIFUS neuromodulation group after the 24-hour follow-up time point^37^. Both patients had histories of similar mood swings, and their adverse symptoms resolved within two weeks. In Cheung et al., 4% of subjects reported headache, and one subject reported vomiting; the reported symptoms subsided within 2 hours^39^.

**Table 2.** Summary of findings.

| LIFUS neuromodulation as a treatment for MDD |  |  |  |  |  |
| --- | --- | --- | --- | --- | --- |
| Population: patients with major depressive disorder or animal models showing depressive-like behaviors |  |  |  |  |  |
| Intervention: Low-intensity focused ultrasound neuromodulation |  |  |  |  |  |
| Randomized clinical trials |  |  |  |  |  |
| Outcome | Compared to | Number of subjects<sup>+</sup> | Estimated effect** (95% CI) | Certainty of the evidence<sup>^</sup> (GRADE)<sup>67</sup> | Justification |
| Depression scale score* | Baseline | 82 | -3.369 (-4.905, -1.832) | Moderate | Downgrade 2 certainty level because of heterogeneity and of imprecision<sup>^^</sup>, upgrade 1 level for large effects. |
|  | Sham | 112 | -1.662 (-3.142, -0.182) | Moderate | Downgrade 1 certainty level because of imprecision. |
| Rodent studies |  |  |  |  |  |
| Sucrose preference index | Sham | 172 | 1.288 (0.864, 1.713) | Low | Downgrade 1 certainty level because of high heterogeneity, upgrade 1 |
|  |  |  |  |  | level because of large effects |
| Immobility time in the forced swimming test | Sham | 192 | -0.913 (-1.567, -0.259) | Very low | Downgrade 2 certainty level because of high heterogeneity and imprecision. |
| Immobility time in the tail suspension test | Sham | 123 | -1.851 (-2.672, -1.030) | Low | Downgrade 1 certainty level because of high heterogeneity, upgrade 1 level because of large effects |

<sup>†</sup>Number of subjects refers to the intent-to-treat (ITT) population. Subjects without available ITT data were excluded.
\*includes Hamilton Depression Rating Scale and the Montgomery-Åsberg Depression Rating Scale
\*\*measured by Cohen's d
<sup>^</sup>For preclinical study, the initial certainty level of the evidence starts is low due to risk of bias. For randomized controlled trials, the initial certainty level of the evidence is high.
<sup>^^</sup> We downgrade the certainty of evidence due to imprecision if either of the following conditions was met: (1) the optimal information size (OIS) criterion was not fulfilled, assuming the clinical meaningful effect of 0.5 Cohen's d; or (2) the 95% confidence interval for the effect size overlapped with the clinical meaningful effect.

## Discussion

This systematic review critically analyzed published studies reporting the use of ultrasound to treat major depressive disorder and depressive-like behaviors in humans and rodent models. The application techniques included magnetic resonance–guided focused ultrasound for anterior capsulotomy and low-intensity transcranial ultrasound for neuromodulation. The current literature suggests a responder rate of 53.85% for MRgFUS capsulotomy. The estimated odds ratio for LIFUS neuromodulation is 2.8 (69.2% response rate in the treatment group and 44.2% for sham control), compared to 1.1 – 6.0 for transcranial magnetic stimulation, 1.1 for invasive vagus nerve stimulation, 5.5 for deep brain stimulation, 1.8 for transcutaneous vagus nerve stimulation, 2.7–8.9 for electroconvulsive therapy, and 1.6–2.3 for transcranial direct current stimulation (Please see **Table 3** for details). LIFUS had a large effect on mitigating depressive-like behaviors in rodent studies, suggesting its promising translation in humans. Neuromodulation strategies for psychiatric disorders are evolving rapidly. For example, optimized protocols and personalized targeting are expected to improve TMS efficacy^78^. To bring ultrasound therapy for MDD in line with clinical progress achieved by established neuromodulation therapy, several key challenges need to be addressed. To date, there are 11 completed or ongoing clinical trials registered on ClinicalTrials.gov that investigate ultrasonic interventions in treating MDD (NCT06085950, NCT05697172, NCT06285474, NCT03421574, NCT06320028, NCT06013384, NCT05301036, NCT04405791, NCT05551585,NCT02348411, NCT05006365). The limited number of clinical studies and small patient cohorts is a limitation of this review. This highlights the need for larger trials to establish the clinical efficacy and safety profiles of HIFUS and LIFUS for treating depression.

**Table 3.**
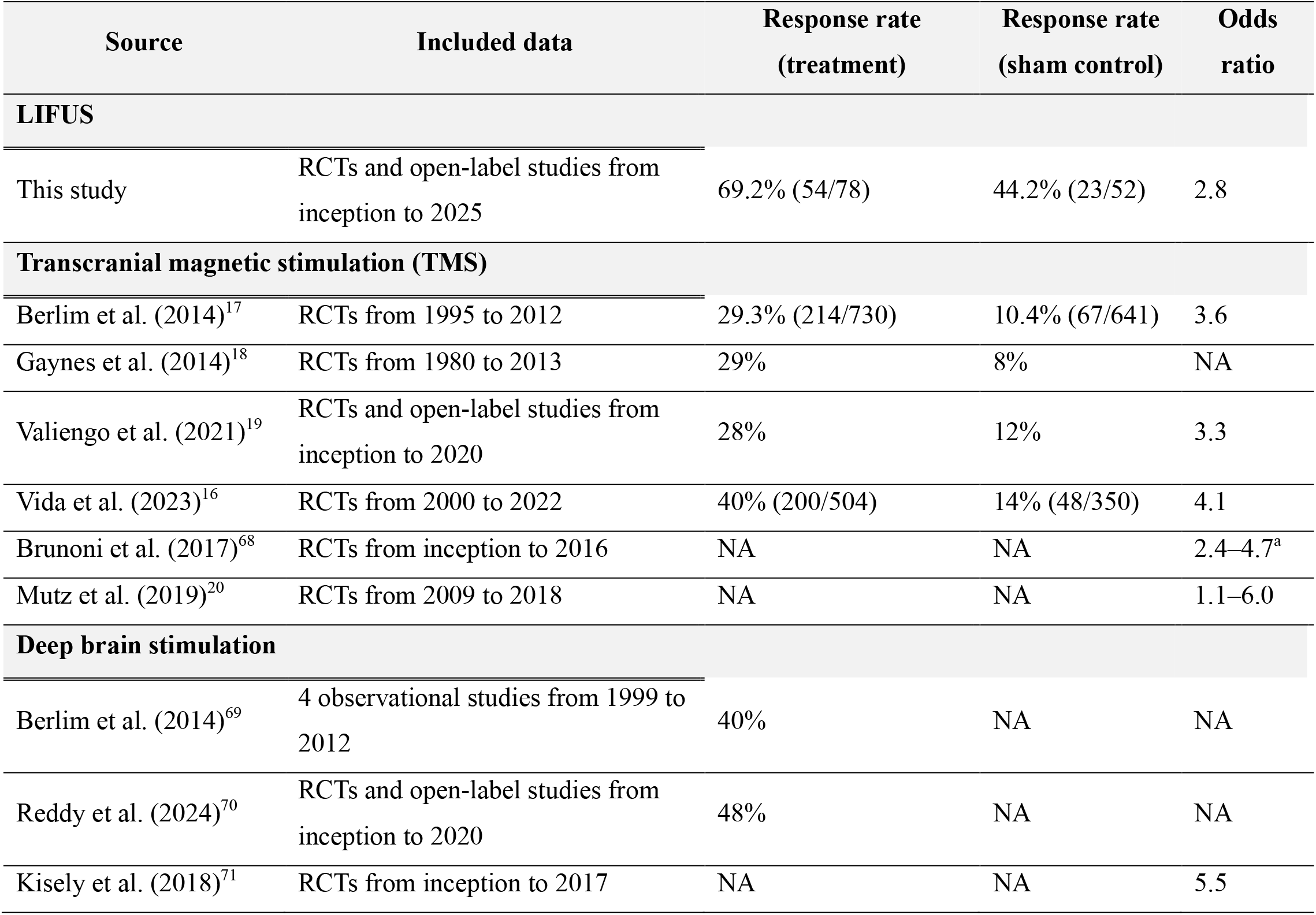
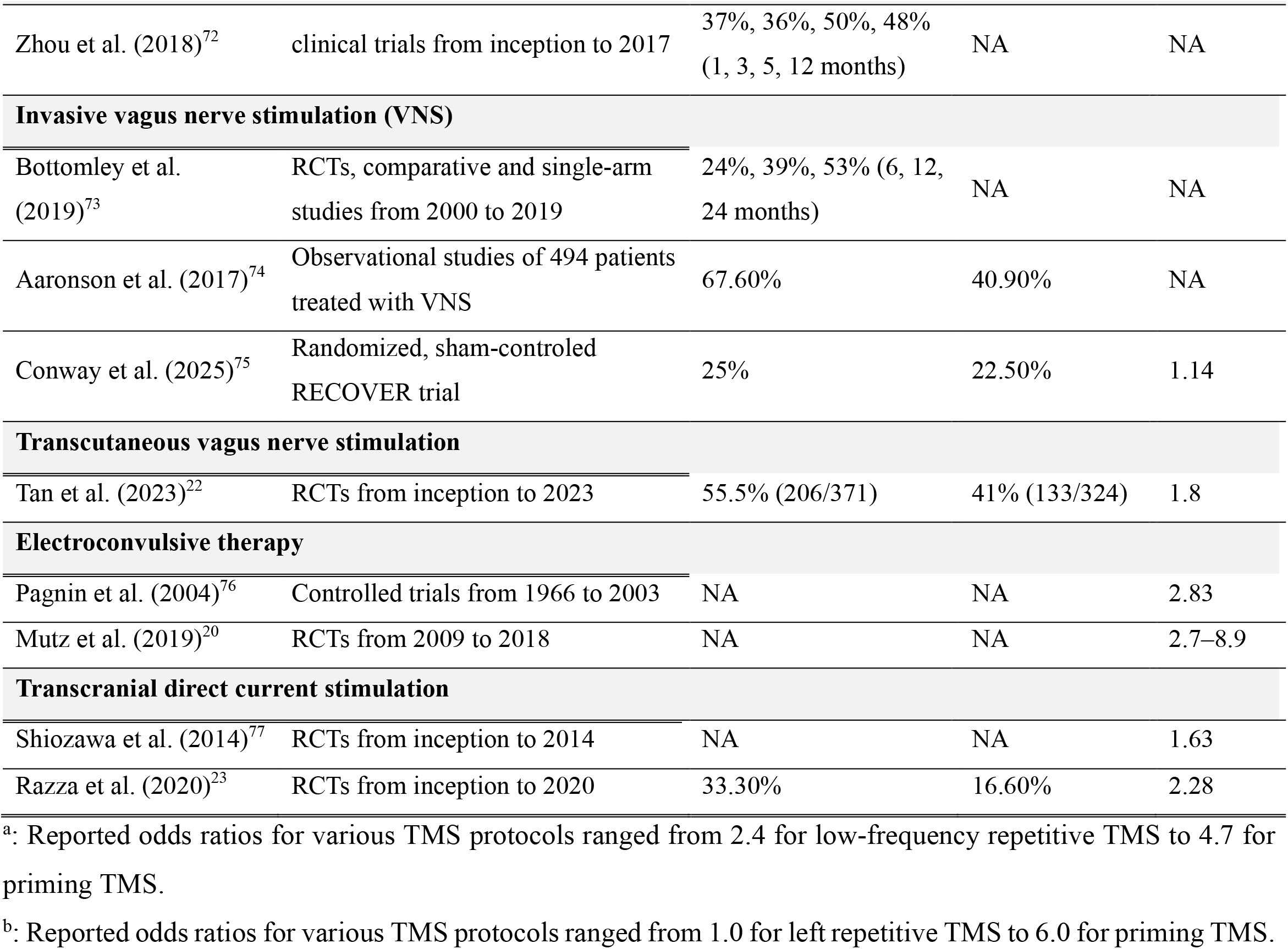
Key systematic reviews and studies reporting clinical efficacy of neuromodulation treatments for depression. This table summarizes high-quality, relevant systematic reviews and key studies selected based on methodological rigor, comprehensiveness, and scientific impact. RCT = randomized controlled trial. Response is defined as ≥50% reduction in the depression rating scale score. When available, we report response rates for the treatment and sham control groups and odds ratios. Odds ratios compare the odds of response in the treatment group to those in the sham control group. Higher odds ratios favor the treatment. NA indicates data not available.

| Source | Included data | Response rate<br>(treatment) | Response rate<br>(sham control) | Odds<br>ratio |
| --- | --- | --- | --- | --- |
| **LIFUS** |  |  |  |  |
| This study | RCTs and open-label studies from inception to 2025 | 69.2% (54/78) | 44.2% (23/52) | 2.8 |
| **Transcranial magnetic stimulation (TMS)** |  |  |  |  |
| Berlim et al. (2014)<sup>17</sup> | RCTs from 1995 to 2012 | 29.3% (214/730) | 10.4% (67/641) | 3.6 |
| Gaynes et al. (2014)<sup>18</sup> | RCTs from 1980 to 2013 | 29% | 8% | NA |
| Valiengo et al. (2021)<sup>19</sup> | RCTs and open-label studies from inception to 2020 | 28% | 12% | 3.3 |
| Vida et al. (2023)<sup>16</sup> | RCTs from 2000 to 2022 | 40% (200/504) | 14% (48/350) | 4.1 |
| Brunoni et al. (2017)<sup>68</sup> | RCTs from inception to 2016 | NA | NA | 2.4–4.7<sup>a</sup> |
| Mutz et al. (2019)<sup>20</sup> | RCTs from 2009 to 2018 | NA | NA | 1.1–6.0 |
| **Deep brain stimulation** |  |  |  |  |
| Berlim et al. (2014)<sup>69</sup> | 4 observational studies from 1999 to 2012 | 40% | NA | NA |
| Reddy et al. (2024)<sup>70</sup> | RCTs and open-label studies from inception to 2020 | 48% | NA | NA |
| Kisely et al. (2018)<sup>71</sup> | RCTs from inception to 2017 | NA | NA | 5.5 |
| Zhou et al. (2018)<sup>72</sup> | clinical trials from inception to 2017 | 37%, 36%, 50%, 48%<br>(1, 3, 5, 12 months) | NA | NA |
| **Invasive vagus nerve stimulation (VNS)** |  |  |  |  |
| Bottomley et al. (2019)<sup>73</sup> | RCTs, comparative and single-arm studies from 2000 to 2019 | 24%, 39%, 53% (6, 12, 24 months) | NA | NA |
| Aaronson et al. (2017)<sup>74</sup> | Observational studies of 494 patients treated with VNS | 67.60% | 40.90% | NA |
| Conway et al. (2025)<sup>75</sup> | Randomized, sham-controlled RECOVER trial | 25% | 22.50% | 1.14 |
| **Transcutaneous vagus nerve stimulation** |  |  |  |  |
| Tan et al. (2023)<sup>22</sup> | RCTs from inception to 2023 | 55.5% (206/371) | 41% (133/324) | 1.8 |
| **Electroconvulsive therapy** |  |  |  |  |
| Pagnin et al. (2004)<sup>76</sup> | Controlled trials from 1966 to 2003 | NA | NA | 2.83 |
| Mutz et al. (2019)<sup>20</sup> | RCTs from 2009 to 2018 | NA | NA | 2.7–8.9 |
| **Transcranial direct current stimulation** |  |  |  |  |
| Shiozawa et al. (2014)<sup>77</sup> | RCTs from inception to 2014 | NA | NA | 1.63 |
| Razza et al. (2020)<sup>23</sup> | RCTs from inception to 2020 | 33.30% | 16.60% | 2.28 |

<sup>a</sup>: Reported odds ratios for various TMS protocols ranged from 2.4 for low-frequency repetitive TMS to 4.7 for priming TMS.
<sup>b</sup>: Reported odds ratios for various TMS protocols ranged from 1.0 for left repetitive TMS to 6.0 for priming TMS.

### Challenges and future directions for HIFUS

A major challenge in the development of HIFUS for thermal ablation is its inconsistent success rate. Although MRgFUS is noninvasive and has minimal reported serious adverse effects, it does not consistently achieve successful ablation of the anterior limb of the internal capsule for treating MDD. MRgFUS efficacy in MDD patients with successful lesioning is comparable to that of traditional bilateral anterior capsulotomy, which has a reported success rate of approximately 40%^79^. Anterior capsulotomy is the most effective and safest option for ablative interventions in MDD compared with other targets in ablative surgery, such as bilateral anterior cingulotomy and bilateral subcaudate tractotomy^79^. Therefore, exploring alternative ablative targets for MRgFUS is unlikely to yield substantial improvements in clinical outcomes. Instead, efforts should focus on refining patient selection through the use of neuroimaging or neurophysiological biomarkers that predict treatment response^80^. Key predictors of successful ablation, including skull density ratio and skull thickness, can be integrated with these biomarkers to identify candidates most likely to benefit from ultrasound-based interventions. Furthermore, individualized functional targeting, applied in other brain stimulation techniques such as transcranial magnetic stimulation, may offer a model for tailoring MRgFUS treatments to the unique neurophysiological profiles of individual patients^81^.

### Challenges and future directions for LIFUS

The application of LIFUS therapy for patients with MDD faces two primary challenges: limited exploration of its parameter space and an incomplete understanding of its underlying mechanisms. All studies included in this review were published after 2017, which may explain the lack of comprehensive investigations into the effects of specific parameters. We anticipate that future research will expand this parameter space, as LIFUS has a medium-to-strong effect on reducing depressive behaviors in animal models and human clinical studies. A further challenge is the lack of rapid and reliable biomarkers of mood. The therapeutic effects of ultrasound in MDD are typically evaluated with outcome measures that unfold over weeks, which limits a systematic and comprehensive exploration of the parameter space. Structural and functional neuroimaging have been investigated as potential ways to obtain shorter-term feedback, although no definitive biomarkers have yet been established^82^.

No formal guidelines currently exist to determine the optimal ultrasound stimulation sites for treating depression. Several brain regions and peripheral targets have been explored, and the current literature contains considerable variations in treatment protocols. It would be useful to establish whether a universal stimulation target exists or if individualized approaches will ultimately prove more effective. Despite variations in treatment protocol, LIFUS consistently reduced depressive-like behaviors in rodent studies and mitigated depressive symptoms in human clinical trials. However, the reported effects of LIFUS in clinical trials were generally transient, lasting from one week to five weeks. To better characterize the time course of LIFUS efficacy, we recommend that future studies include follow-up assessments after the immediate post-treatment period. Depression is increasingly conceptualized as arising from dysfunctional connectivity across distributed neural circuits. Directing LIFUS to key nodes within these aberrant networks may disrupt pathological activity and yield therapeutic benefits^83^. This hypothesis warrants further investigation.

The precise mechanisms driving LIFUS neuromodulation have not been established. It will be crucial to determine whether these mechanisms are mediated by thermal, mechanical, or cavitation-based effects, or a combination of these effects, to optimize ultrasound stimulation protocols and align them with established neurobiological pathways of depression. LIFUS has distinct advantages as a noninvasive neuromodulatory technique that penetrates deep brain tissue. Temporal interference electrical neurostimulation is a potential rival technique that uses multiple high-frequency electric fields to provide steerable and focal stimulation^84^. How small a focal volume may be achieved via temporal interference stimulation remains an open question, but an animal study showed that temporal interference stimulation can activate the mouse hippocampus without stimulating the overlying cortex^85^. In comparison, the focal volume for transcranial FUS is 23 mm^3^ under a 0.5 mm mouse skull, a size comparable to the volume of the mouse hippocampus^86^. Although both approaches can noninvasively stimulate subcortical structures, their differing modes of action (electrical vs mechanical) may differentially confer greater efficacy in specific patient populations or disease states. Future research should systematically compare the neurophysiological outcomes of ultrasound neuromodulation and temporal interference across patient populations. These studies may identify disease categories that respond better to mechanical or thermal mechanisms.

## Limitations

Our systematic review has limitations. We evaluate the efficacy of MRgFUS anterior capsulotomy in MDD patients using per-protocol estimates because intention-to-treat (ITT) data were not reported in the included studies. A meta-epidemiological study suggests that per-protocol analyses, on average, yield treatment effects approximately 2% higher than ITT analyses^87^. Our analysis based on per-protocol estimates may overstate the true treatment effect. Another limitation concerns the heterogeneity in stimulation targets across the included LIFUS clinical trials and preclinical studies. Two targets were used in LIFUS clinical trials: the dorsolateral prefrontal cortex (PFC) and the subcallosal cingulate cortex. We included studies targeting the dorsolateral PFC or the subcallosal cingulate cortex in our meta-analysis. In fact, depression is becoming increasingly recognized as a network disorder^88,89^. The left dorsolateral PFC and the subcallosal cingulate have received the most attention due to the consistency of their depression-related abnormalities^5,89^. In addition, recent studies indicated dorsolateral PFC and subcallosal cingulate are functionally connected and parts of a circuit implicated in the pathophysiology of depression^5^. Changes in functional connectivity between the anterior cingulate and left dorsolateral PFC predict the treatment response of PFC stimulation^8^. As a secondary analysis, we evaluated the response rate by target. The synthesis response rate was 68.9% in the treatment group and 47.6% in the sham group for LIFUS targeting PFC. The synthesis response rate was 70.6% in the treatment group and 30.0% in the sham group for LIFUS targeting the subcallosal cingulate cortex. In preclinical LIFUS studies, there is greater heterogeneity in the stimulation target. The pooled effect sizes across these studies should be interpreted with consideration of the variability in anatomical targets and the mechanistic pathways engaged. Given the absence of a consensus on the optimal stimulation target for LIFUS for treating depression, we did not consider target variability as a deviation from the intended intervention when assessing risk of bias using RoB 2. To further advance the development of therapeutic ultrasound in treating depression, future systematic reviews should aim to conduct target-stratified meta-analyses as more data accumulate. Once an optimal stimulation target is established, target deviations should also be incorporated into risk of bias assessments.

The response rate in the sham control group was relatively high (44.2%), raising concerns about the extent to which nonspecific factors may have contributed to clinical improvement. In Li et al., the sham group received 30 minutes of TMS intervention, while the treatment group underwent 15 minutes of TMS treatment and 15 minutes of LIFUS treatment^34^. After excluding this study, the synthesized sham response rate dropped to 18.2%, which aligns more closely with sham response rates reported in other neuromodulation modalities (**Table 3**). The relatively high sham response is reminiscent of the substantial placebo effect (13–52% response rate) in antidepressant pharmacotherapy trials^90^. Several factors may contribute to the placebo effect, including longer trial durations, broader public awareness and social acceptance of antidepressant treatment, and recruitment strategies that may attract individuals seeking treatment. A longer trial duration might allow more time for natural recovery or the accumulation of nonspecific therapeutic effects, as well as address patient expectations. These factors likely influence response rates in neuromodulation studies for depression. Therefore, we recommend including sham control conditions whenever feasible to isolate the specific effects of neuromodulation interventions from nonspecific improvements. When sham control is technically challenging, we suggest using comparative control designs, such as waitlist or crossover groups.

## Conclusion

Ultrasound applications open exciting new avenues for treating MDD through noninvasive ablation and neuromodulation. HIFUS for anterior capsulotomy had a response rate of 38.46%, which is comparable to traditional capsulotomy with stereotactic surgery. Future development of HIFUS will benefit from refined patient selection and technological advances to ensure consistent lesioning. LIFUS had a response rate of 69.2% in the treatment group, compared to 44.2% in the sham group in clinical studies. In addition, LIFUS neuromodulation achieved a large effect on resolving depressive-like behavior in rodents and reducing depression scores in humans. However, the mechanism underlying LIFUS neuromodulation has not been established, and the stimulation parameters have not been sufficiently explored. As ultrasound applications continue to evolve, it will be crucial to integrate mechanistic research and refine stimulation protocols to develop a safer and more effective neuromodulation strategy in MDD.

## Supporting information

SYRCLE source data

ROBINS E source data1

ROBINS E source data2

ROBINS E source data3

ROBINS E source data4

## Data Availability

All data produced in the present work are contained in the manuscript

## Data availability

Data supporting the findings of this study are available within the article and its supplementary materials. Analysis codes for meta-analysis are available at https://github.com/GanshengT/Meta_analysis_ultrasound/tree/main.

## Declaration of Generative AI and AI-assisted technologies in the writing process’

During the preparation of this work, the authors used ChatGPT in order to detect grammatical errors. After using this tool, the authors reviewed and edited the content as needed and take full responsibility for the content of the publication.

## Acknowledgments

This work was supported by the National Institutes of Health R01CA276174. The manuscript was edited by the Scientific Editing Service of the Institute of Clinical and Translational Sciences at Washington University, which is supported by an NIH Clinical and Translational Science Award (UL1 TR002345).

## Authorship Statements

All authors designed and conducted the study, including record retrieval, screening, identifying risks of bias and quality assessment. Gansheng Tan extracted and analyzed data and prepared the manuscript draft with important intellectual input from Eric Leuthardt and Hong Chen. All authors reviewed and approved the final version of the manuscript.

## Supporting information

**Supplementary Figure 1.**
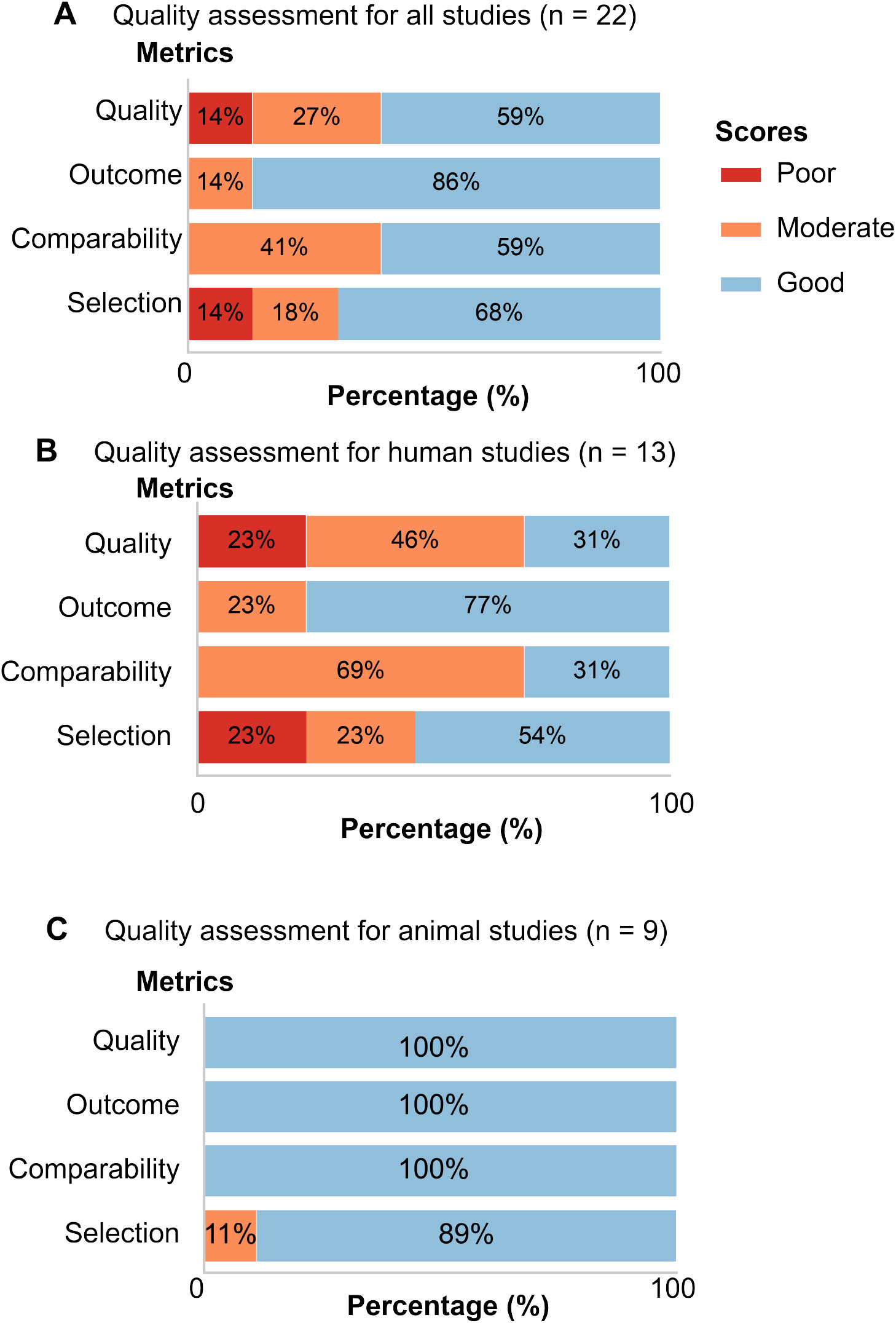
Quality assessment.

## Appendix 1 Quality Assessment

### Quality assessment of studies examining the technique and efficacy of therapeutic ultrasound in the treatment of major depressive disorder

The maximum score for a study is 1 for the Selection, Comparability, and Outcome categories.

#### Selection

1. Representativeness of the exposed cohort
  1: The study includes both genders, and the sample size is> 7. If the study used an animal model, the model should be a valid depression animal model.
  0.5 (somewhat representative): Sample size ≥ 5
  0: case report

#### Comparability

1. Comparability of cohorts in study design and statistical analysis
  1: The study includes both a treatment group and a control group. The outcome measures for both groups are adjusted for baseline measurement.
  0.5: The study outcomes for the treatment group before and after ultrasonic treatment are available. Alternatively, the study includes outcome measures for the treatment group and the control group.
  0: The outcome measures before and after treatment are not comparable, for example, due to different measures used.

#### Outcome

1. Outcome measure
  1: Standard measures or rating scales for depression that have been validated
  0.5: The study includes neuroimaging outcomes, biochemical outcomes, electrophysiological outcomes, or other outcomes that are related to standard rating scales for depression
  0: Subjective report

Thresholds for converting the Quality assessment to AHRQ standards (good, moderate, and poor): Good quality: at least one score for any two of the three categories and at least 0.5 score for the other category Moderate quality: studies that are neither of good quality nor poor quality.

Poor quality: 0 score for any one of the three categories.

## Appendix 2 Meta-analysis using fixed-effect models

In our original protocol, we had pre-specified the use of fixed-effect models based on the assumption of a common true effect across all studies across studies. Given the observed heterogeneity (I^2^ > 50%), this assumption is not met. Therefore, we reported the results of the meta-analysis using random-effects models in the main text. In this appendix, we report the results of the meta-analysis using fixed-effect models for transparency and reference.

In preclinical studies, meta-analysis using fixed-effect models showed that LIFUS had a medium effect (Cohen’s d) on reducing immobility time in the forced swimming test 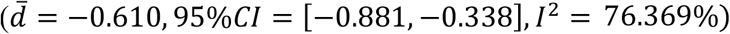. The pooled effect size for the sucrose preference index was large 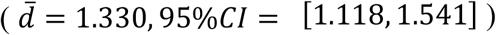. LIFUS reduced immobility time in the tail suspension test with a large effect 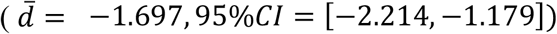. In clinical trials, meta-analysis using fixed-effect models showed that LIFUS had a large effect on reducing depression severity within the ultrasound treatment group compared to the baseline depression score 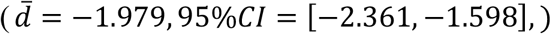. LIFUS also had a large effect on reducing depression severity in the ultrasound treatment group when compared with the sham control group after adjusting for baseline scores 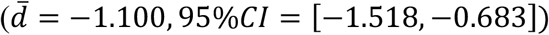.

